# Fully automated detection and differentiation of pandemic and endemic coronaviruses (NL63, 229E, HKU1, OC43 and SARS-CoV-2) on the Hologic Panther Fusion

**DOI:** 10.1101/2020.08.31.20185074

**Authors:** Anne K. Cordes, William M. Rehrauer, Molly A. Accola, Benno Wölk, Birgitta Hilfrich, Albert Heim

## Abstract

The Hologic Panther Fusion (PF) platform provides fully automated CE marked diagnostics for respiratory viruses, including recently SARS-coronavirus 2 (SARS-CoV-2) by a transcription mediated amplification (TMA) assay, but not for the endemic human coronaviruses (hCoV). Therefore, a laboratory developed test (LDT) comprising a multiplexed RT-PCR protocol that detects and differentiates the four hCoV NL63, 229E, HKU1 and OC43 was adapted on the PF.

The novel CE marked Aptima SARS-CoV-2 TMA and the LDT for hCoV were validated with 321 diagnostic specimens from the upper and lower respiratory tract in comparison to two SARS-CoV-2 RT-PCRs (PF E-gene RT-PCR and genesig RT-PCR, 157 specimens) or the R-GENE hCoV / hParaFlu RT-PCR (164 specimens), respectively.

For the endemic hCoV, results were 96.3 % concordant with two specimens discordantly positive in the PF and four specimens discordantly positive in the R-GENE assay. All discordantly positive samples had Ct values between 33 and 39. The PF hCoV LDT identified 23 hCoV positive specimens as NL63, 15 as 229E, 15 as HKU1 and 25 as OC43. The Aptima SARS-CoV-2 TMA gave 99.4 % concordant results compared to the consensus results with a single specimen discordantly positive. Moreover, 36 samples from proficiency testing panels were detected and typed correctly by both novel methods.

In conclusion, the SARS-CoV-2 TMA and the LDT for hCoV enhanced the diagnostic spectrum of the PF for all coronaviruses circulating globally for a multitude of diagnostic materials from the upper and lower respiratory tract.

## Introduction

Human coronaviruses are a common cause for acute respiratory infections of the upper respiratory tract ^1-3^, though occasionally infections of the lower respiratory tract have been described. Especially children, elderly and chronically ill patients are at risk of a more severe and potentially lethal progress of disease ^4-7^. Four different human coronavirusspecies (hCoV 229E, OC43, NL63 and HKU1) are circulating in the human population worldwide. HCoV 229E and OC43 have been known since the 1960s ^8,9^. More than three decades later, in 2004, NL63 ^10^ and in 2005 HKU1 ^11^ were identified. In addition to these hCoVs three zoonotic coronaviruses were discovered: SARS-CoV (causing an outbreak in 2003), MERS-CoV (discovered in 2013 and circulating almost exclusively on the Arabian Peninsula) and SARS-CoV-2 (a novel S*arbecovirus* currently pandemic and probably endemic in future). These cause severe lower respiratory tract infections in humans more frequently ^12,13^. However, many SARS-CoV-2 infections can be asymptomatic or only associated with mild symptoms especially in children and young adults ^14-16^. Moreover, high virus loads were found in presymptomatic patients, which might play a crucial role in virus transmission ^17^.

In acute respiratory infections, the rapid diagnosis of a viral etiology is essential for reducing the amount of prescribed antibiotics and for infection control measures. The introduction of nucleic acid amplification technologies (NAT) such as polymerase chain reaction (PCR) and transcription-mediated amplification (TMA) has been a milestone in the diagnosis of respiratory virus infections in comparison to virus culture and direct fluorescent-antibody assays improving sensitivity. Therefore, NAT is now the gold standard of coronavirus diagnostics ^18,19^. However, respiratory samples were processed in batches usually once every working day for conventional coronavirus PCR diagnostics, thus resulting in long sample to answer times. More recently, fully automatic NAT platforms were developed which process each diagnostic sample individually (“random access”) and thus reduce sample-to-answer time significantly. For example, coronavirus detection can be performed by highly multiplexed, random access PCR platforms ^20-22^ but these cannot be adapted to the individual diagnostics needs of a patient considering his symptoms and the epidemiology of circulating respiratory viruses. For comparison, the Hologic Panther Fusion (PF) is a random access platform that provides a panel of three CE marked and FDA cleared multiplex real-time PCRs that cover influenza virus A/B (Flu A/B), respiratory syncytial virus (RSV), parainfluenza virus 1-4 (ParaFlu), human metapneumovirus (hMPV), adenovirus (AdV) and rhinovirus (RhV) and can be performed according to the individual diagnostic request. These assays were validated for samples from the lower respiratory tract in a previous study ^23^.

Recently, a TMA based assay for SARS-CoV-2 became available for the Panther (Aptima SARS-CoV-2 assay) ^24^ which is CE marked and FDA cleared for emergency use (EUA) for samples from the upper respiratory tract (nasopharyngeal swabs, washes and aspirates; nasal swabs and aspirates, oropharyngeal swabs). In addition, an early laboratory developed real time PCR for the SARS-CoV E-gene ^25^ and a FDA EUA cleared RT-PCR for the SARS-CoV-2 Orf1ab which is only available in the U.S. ^26^, can be applied on the PF platform. However, the endemic human coronaviruses were not yet covered. As these are responsible for about 13-19.7 % of the respiratory infections ^2,27^, we used the PF Open Access tool to establish and validate a multiplexed laboratory developed test (LDT) that detects and differentiates the four endemic hCoVs 229E, OC43, NL63 and HKU1.

## Material and Methods

### Proficiency testing panels

36 proficiency testing specimens provided by Instand (Düsseldorf, Germany) in the years 2017-2020 (sample numbers 340029 – 340050 and 340052 - 340065) were tested with the novel LDT for endemic hCoV NL63, 229E, HKU1, OC43 and the Aptima SARS-CoV-2 assay.

### Panels of diagnostic specimens

In total, 321 patient samples from the upper (URT) and lower respiratory tract (LRT) were included in the panel.

164 patient samples (91 bronchial alveolar lavages (BAL), 41 nasopharyngeal swabs (NPS), 20 pharyngeal lavages (PL), 6 bronchial lavages (BL), 4 tracheal secretions (TS), 2 nasal swabs (NS)) with a diagnostic request for coronavirus were tested by the hCoV / hParaFlu R-GENE (Biomerieux, Marcy-l’Étoile, France) as the routine diagnostic procedure and the PF LDT. Specimens originated between 03/2017 and 02/2020.

For validation of the Aptima SARS-CoV-2 TMA, 157 specimens (8 BAL, 125 NPS, 1 PL, 7 BL, 3 TS, 6 NS and 7 tracheal swabs (TRS)) with a diagnostic request for SARS-CoV-2 were tested in the PF E-gene RT-PCR ^25^ as the routine diagnostic procedure and subsequently in the Aptima TMA and the genesig RT-PCR (Primerdesign, Chandler’s Ford, UK).

### Ethical statement

All 321 diagnostic samples originated from patients who had agreed to the anonymised use of their clinical data at hospital admission (informed consent). As specimens were tested according to diagnostic requests only and medical data were anonymized and analyzed only retrospectively, ethical approval is not required in Germany (confirmed by e-mails of the local ethical committee “Ethikkommission der Medizinischen Hochschule Hannover”, 19th of September, 2019 and 6th of May 2020).

### Sample preparation

Swabs were resuspended in 1.5 ml PBS^-^. Liquid samples (lavages and secretions) were loaded without prior processing of the sample. However, viscous samples were diluted 1:2 in PBS^-^ to avoid clotting in the pipette tip. For testing on the PF, 500 µl of the specimen (patient specimen and proficiency testing samples) were transferred into a Specimen Lysis Tube (SLT) that contained 710 µl Specimen Transport Media (STM). For all assays that were performed on the PF, 360 µl of this sample were pipetted automatically for nucleic acid extraction.

### Coronavirus (NL63, 229E, HKU1 and OC43) LDT

SLTs were automatically processed on the Panther Fusion system including nucleic acid extraction, reverse transcription and real time PCR using the Open Access RNA/DNA enzyme cartridge, the Extraction Reagent-S, the Capture Reagent-S, the Enhancer Reagent-S, the internal control and the primer and probe recon solution (PPR). The PPR (table 1) contained coronavirus specific (targeting the respective N gene) as well as internal control (IC) primers and probes, MgCl_2_, KCl, Tris buffer and nuclease free water (all ThermoFisher, Waltham, MA, USA). Coronavirus specific primer and probe sequences were published recently ^28^ but probes were labelled with different fluorescent dyes and quenchers (supplementary table 1). Moreover, a second internal quencher (ZEN) was added to the 229E specific probe and the NL63 specific probe. Stability of the PPR on the Panther Fusion was validated for a period of 14 days (supplementary table 2).

**Table 1:**
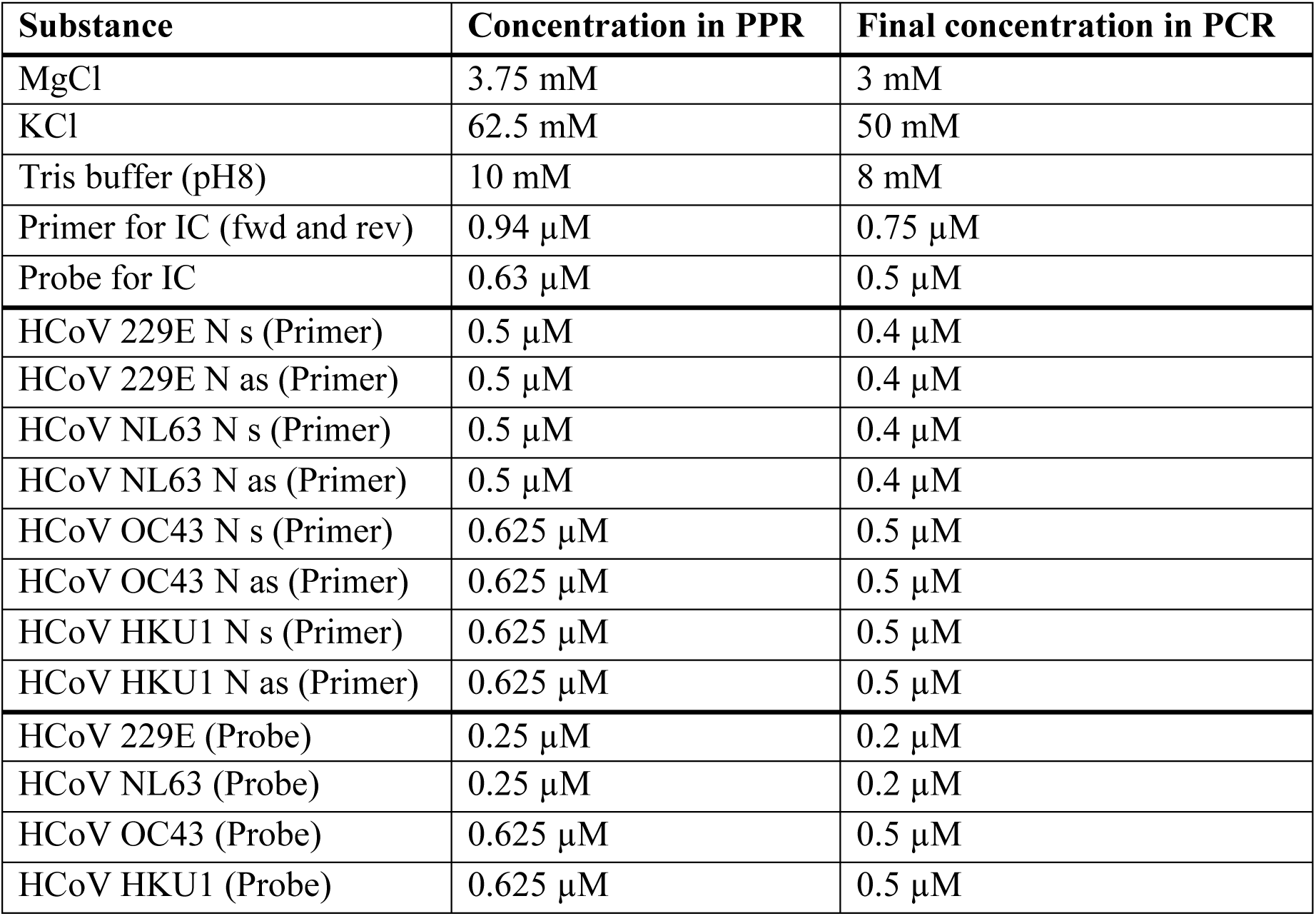
Concentrations in PPR and final concentration in the PCR reaction. The concentration of the components in the PPR is higher by 1.25 x than in the final PCR reaction.

Settings in the PF software for automatic processing were: extraction volume 360 µl and “sample aspiration height” low. The reverse transcription was performed at 50 °C for 8:28 minutes followed by the activation of the Taq polymerase at 95 °C for 2:00 minutes. The two step thermocycling protocol consisted of 45 cycles each with a denaturation at 95 °C for 5 seconds and the elongation and fluorescence detection at 60 °C for 22 seconds. Fluorescence analysis settings in the PF software were set as following: The analysis start cycle was set at 10, the baseline correction was enabled and the slope limit was set at 25. As a positivity criteria the Ct threshold in the channels 1 (FAM for 229E), 2 (HEX for NL63) and 3 (ROX for HKU1) was set to 200 RFU, in channel 4 (Quasar 670 for OC43) to 300 RFU and in channel 5 (Quasar 705 for IC) to 1000 RFU. The crosstalk correction was set to 1 % for the combination of emitter channel 1 and receiver channel 2, as well as the combination of emitter channel 5 and receiver channel 4. A result was defined as valid if at least one positive result in channels 1-5 was detected.

### HCoV/hParaFlu R-GENE RT-PCR

Nucleic acid extraction was performed from 200 µl sample on a QIAcube using the DNeasy blood kit (QIAGEN, Hilden, Germany) which has been previously validated for this purpose^23^. Of the eluate, 10 µl were tested by the hCoV / hParaFlu R-GENE assay (bioMérieux, Marcy-l’Étoile, France) on an ABI 7500 sequence detection system (ABI, Foster City, CA, U.S.). This assay has been validated previously for multiple types of respiratory specimens.

### Aptima SARS-CoV-2 TMA

SLTs were loaded on the Panther Fusion and the assay was performed automatically as described in the package insert ^24^.

### SARS-CoV-2 E gene RT-PCR

The SARS-CoV-2 E gene RT-PCR was performed with the Open Access tool on the Panther Fusion as described previously ^25^.

### Genesig SARS-CoV-2 RT-PCR

Nucleic acid extraction was performed from 200 µl sample on a QIAcube using the DNeasy blood kit (QIAGEN, Hilden, Germany). Of the eluate, 10 µL were tested by the genesig coronavirus COVID-19 Real-Time PCR assay (Primerdesign, Chandler’s Ford, UK) on an ABI 7500 sequence detection system.

### Amplification efficiency and evaluation of LOD

The amplification efficiency was calculated from serial dilutions of reference samples if available. Supernatants from cell cultures, positive for 229E, NL63 and OC43 were provided by the national reference centre for coronaviruses at Charité, Berlin. For SARS-CoV-2, the quantified quality control material AccuPlex SARS-CoV-2 Verification Panel (cat # 0505-0129 from Seracare, Milford, MA) was used. For HKU1, no cell culture supernatant was available. Therefore, a patient sample that had been tested highly positive in the hCoV / ParaFlu R-GENE assay was used for estimating the amplification efficiency.

For the Aptima SARS-CoV-2, the limit of detection (LOD) was determined from half logarithmic serial dilutions.

### Data analysis and statistics

The amplification efficiency was calculated with the following formula: Amplification efficiency (E) = 10^−1/m^ (with m being the slope of the amplification curve). For the Aptima SARS-CoV-2 TMA assay the LOD (95% probability of detection according to 2002/364/EC) was calculated by probit analysis from 27 replicates of a dilution series of the Accuplex standard using the statistics program SSPS (Version 15.0). For panels of diagnostic specimens, concordances of assays and positive percent agreement (PPA, “sensitivity”) and negative percent agreement (NPA, “specificity”) of PF assays were calculated. To evaluate the clinical performance of the Aptima SARS-CoV-2 TMA compared to those of the two other RT-PCRs (Panther Fusion E-gene RT-PCR and genesig COVID-19 RT-PCR), a consensus result was defined as a concordant result in at least two of three tests.

## Results

### Proficiency testing panel specimens and cross-reactivity

36 proficiency testing specimens provided by Instand (Düsseldorf, Germany) were tested with the hCoV LDT and Aptima SARS-CoV-2 TMA. All samples were detected correctly. From these samples, 31 were coronavirus positive samples (10 OC43, 3 NL63 and 4 229E, 4 SARS-CoV-2, 10 MERS-CoV) and 5 were negative for these coronaviruses. As anticipated, MERS-CoV positive specimens were negative in the hCoV LDT and the Aptima SARS-CoV-2 TMA. Hence, cross-reactivity with MERS-CoV could be excluded. Furthermore, an inactivated SARS-CoV positive cell culture supernatant originating from 2003 gave a negative result in the Aptima SARS-CoV-2 TMA.

### Comparison of test performance for coronavirus NL63, 229E, HKU1 and OC43 detection

A panel of 164 archived diagnostic specimens was tested with the hCoV LDT on the Panther Fusion and results were compared to the initial results of the hCoV / ParaFlu R-GENE assay (table 2). Overall, the concordance of results was 96.3 %. The positive percent agreement (PPA, “sensitivity”) of the LDT on the Panther Fusion was determined as 94.9 % and the negative percent agreement (NPA, “specificity”) as 97.6 % in relation to the R-GENE assay. Samples with discordant results had Ct values of 33, 34, 36 and 39 in the R-GENE assay and 38 (two samples) in the PF LDT. In contrast to the R-GENE assay, the PF LDT provided differentiation of hCoV samples: 25 of 77 were identified as OC43, 23 as NL63, 15 as 229E and 15 as HKU1. Among these, one sample was co-infected with 229E (with a Ct value of 21) and OC43 (Ct value 32).

**Table 2:** Comparison of the PF LDT and the R-GENE hCoV results

| **Diagnostic specimens**<br>(from URT and LRT) | **R-GENE positive** | **R-GENE negative** | **Total** |
| --- | --- | --- | --- |
| **PF LDT positive** | 75 | 2 | 77 |
| **PF LDT negative** | 4 | 83 | 87 |
| **Total** | 79 | 85 | 164 |

### Diagnostic performance of the Aptima SARS-CoV-2 TMA

Of 157 samples tested, 56 were found to be consensus positive for SARS-CoV-2 RNA in any two of the tree methods (table 3). Thus, the PPA of the Aptima SARS-CoV-2 TMA and of the PF E-gene RT-PCR were determined as 100.0 % and the NPA as 99.0 %, whereas the genesig COVID-19 PCR had a PPA of 87.5 % and a NPA of 100 % (table 3). In the SARS-CoV-2 E-gene RT-PCR the median Ct of positive samples was 34.2 (range 17.5 – 40.8) with an interquartile range of 6.4 and in the genesig RT-PCR the median Ct was 32.7 (range 19.2 – 39.2) and the inter quartile range 6.4. Thus the Aptima SARS-CoV-2 TMA detected multiple faintly positive samples correctly. Moreover, 23 of 157 samples originated from the lower respiratory tract and were all detected correctly in the Aptima in comparison to the consensus result with 15 of these positive for SARS-CoV-2 RNA. Detailed results for various diagnostic materials are presented in table 4.

**Table 3.**
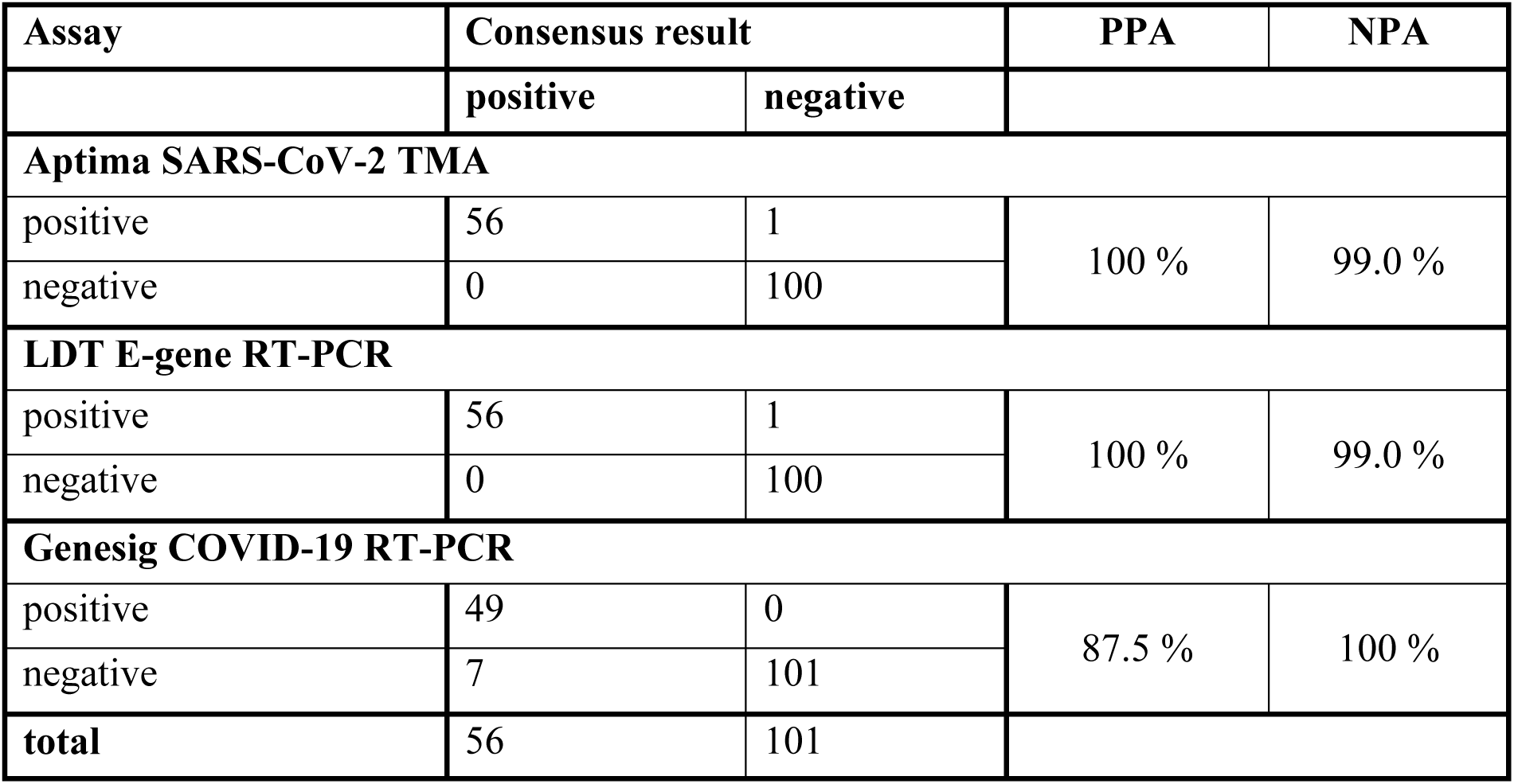
Comparison of three SARS-CoV-2 NATs to the consensus results.

**Table 4.** Performance of SARS-CoV-2 assays with 157 specimens from the upper and lower respiratory tract. Samples that were found to be positive in at least two out of three assays were declared as consensus positive. For negative samples the criteria was implemented respectively.

| **Material\*** | **Aptima SARS-CoV-2** |  | **E-gene LDT RT-PCR** |  | **Genesig RT-PCR** |  |
| --- | --- | --- | --- | --- | --- | --- |
|  | positive of consensus positive | negative of consensus negative | positive of consensus positive | negative of consensus negative | positive of consensus positive | negative of consensus negative |
| **BAL** | 5/5 | 3/3 | 5/5 | 3/3 | 4/5 | 3/3 |
| **BL** | 5/5 | 2/2 | 5/5 | 2/2 | 4/5 | 2/3 |
| **NS** | 3/3 | 3/3 | 3/3 | 3/3 | 3/3 | 3/3 |
| **NPS** | 38/38 | 86/87 | 38/38 | 86/87 | 34/38 | 87/87 |
| **PL** | 0/0 | 1/1 | 0/0 | 1/1 | 0/0 | 1/1 |
| **TRS** | 3/3 | 4/4 | 3/3 | 4/4 | 2/3 | 4/4 |
| **TS** | 2/2 | 1/1 | 2/2 | 1/1 | 2/2 | 1/1 |

\*bronchial alveolar lavages (BAL), bronchial lavages (BL), nasal swabs (NS), nasopharyngeal swabs (NPS), pharyngeal lavages (PL), tracheal swabs (TRS), tracheal secretions (TS)

### Amplification efficiencies and LOD

The LOD of the Aptima SARS-CoV-2 TMA was 288 copies/ml (95 % confidence interval 191 – 755 copies/ml) determined as the 95 % probability of detection. In case of 229E, NL63, OC43 and HKU1 the LODs were not determined due to the limited availability of quantified reference materials but amplification efficiencies of PCR were calculated. Amplification efficiencies for 229E, NL63, OC43 and HKU1 were 1.89, 1.92, 2.0 and 1.95, respectively (figure 1). For SARS-CoV-2 an amplification efficiency cannot be calculated because this assay uses TMA technology with subsequent hybridization to a SARS-CoV-2 specific probe.

**Figure 1:**
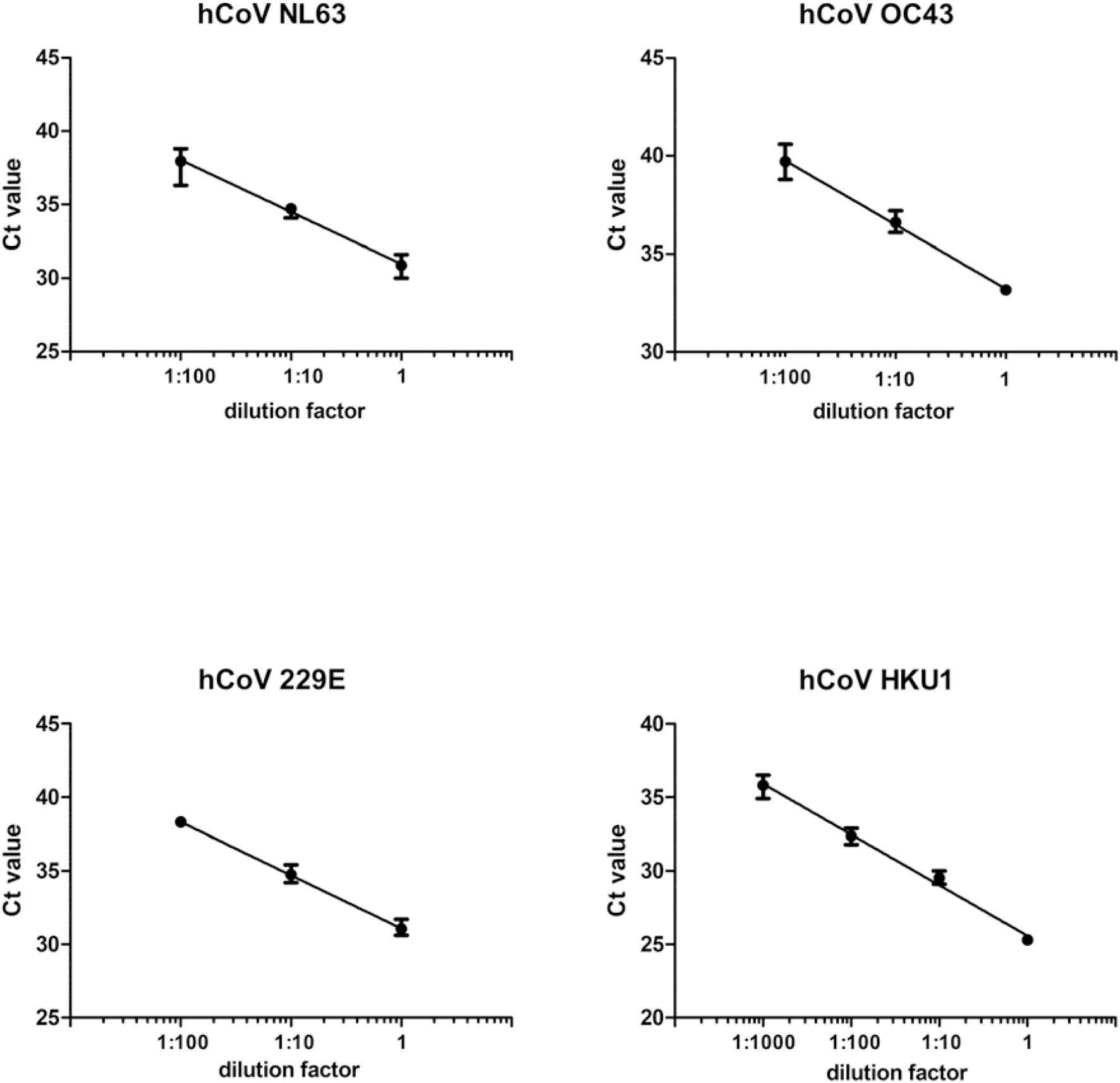
Amplification efficiency. For NL63, OC43 and 229E reference material from cell culture supernatants was applied. For the dilution series of HKU1, a highly positive diagnostic specimen was used. Each dot represents the mean out of three individual test results. The error bars display the range of Ct values.

## Discussion

Rapid diagnostics of respiratory virus infections is essential in order to reduce the amount of prescribed antibiotics, as well as for complying hygiene regulations. Identification of the virus gives the medical staff information on whether the patient should be isolated and which personal protective equipment should be applied. To optimise the sample-to-answer time, the PF platform provides random access testing instead of batch-wise testing as in the hCoV / hParaFlu R-GENE assay and the genesig COVID-19 RT-PCR. Previously, only assays for influenza virus A/B, respiratory syncytial virus, parainfluenza virus 1-4, human metapneumovirus, adenovirus and rhinovirus were available for the PF. Due to the SARS-CoV-2 pandemic, a RT-PCR for the E-gene of SARS-CoV-2 was rapidly established for the open access capability of the PF ^25^. Recently, a CE marked and FDA cleared SARS-CoV-2 TMA, which provides dual target detection and a high analytical sensitivity (low LOD) became available (Aptima SARS-CoV-2) ^29,30^. Even though the mutation rate of SARS-CoV-2 seems to be low to moderate ^31^, single nucleotide mutations localized at a primer binding site in a PCR targeting the E gene have been described that subsequently could give false negative results ^32^. Therefore, dual target detection should be preferred over single-target assays in SARS-CoV-2 diagnostics to preclude false negative results due to mutations affecting the primer or probe binding site.

We evaluated the Aptima SARS-CoV-2 assay with a panel of 157 archived diagnostic specimens which included a multitude of diagnostic materials from the upper and lower respiratory tract. Although the intended use of the Aptima SARS-CoV-2 is limited to diagnostic materials from the upper respiratory tract, SARS-CoV-2 RNA was detected with 100 % PPA and 99.0 % NPA. These results compared favorably to other studies, which had only included nasopharyngeal swabs (94.7 % to 100 % PPA and 98.7 % to 100 % NPA) ^29,30,33^. However, detection of SARS-CoV-2 in materials from the LRT is crucial as it was shown that nasopharyngeal swabs may turn negative in the course of infection prior to specimens from the LRT ^16,34-36^.

Only a single sample (NPS) gave a discordant (false positive) result in the Aptima SARS-CoV-2 assay in comparison to the consensus results. However, this sample was a follow up sample from a convalescent patient who was diagnosed as SARS-CoV-2 RNA positive in previous samples. Therefore, the Aptima SARS-CoV-2 assay may have still detected residual SARS-CoV-2 RNA due to its very low LOD. In this study, the LOD was determined to be 288 copies/ml, whereas others had a 100 % detection rate at 83 copies/ml also using quality control material from Seracare but with only 20 replicates tested ^30^. Nevertheless, the LOD of the Aptima SARS-CoV-2 assay was even lower than the LOD of the E-gene RT-PCR on the PF (315 copies/ml). Both assays run automatized on the PF from samples that have been processed identically before having been loaded onto the platform. On the PF, 360 µl of the sample are pipetted for both assays, however, only 10 % of the extracted nucleic acid is used for the PCR reaction, while in the TMA the total volume is applied. Although highly sensitive, the Aptima SARS-CoV-2 assay is highly specific and does not cross-react SARS-CoV (of 2003), MERS-CoV and the endemic hCoV ^30^, which was confirmed in our study by testing proficiency panel specimens. Previously, cross reactivity with other respiratory viruses was excluded extensively ^30^.

Diagnosis of endemic hCoV infections is not only required in URT infections but also in more severe LRT infections as e.g. bronchiolitis ^5-7^. For example, hCoV NL63 binds to the same cellular receptor as SARS-CoV-2 ^37^ and can be associated with severe LRT infections ^10,38^. Therefore, rapid diagnosis of hCoV infections is essential and can be achieved with the novel hCoV LDT on the PF. Furthermore, a potential cross immunity to SARS-CoV-2 infection following infection with hCoV was suggested ^39^. Therefore, detection and classification of hCoV might be of importance in the future, to predict the prognosis of SARS-CoV-2 infection. The analysis of 164 diagnostic hCoV specimens from the upper and lower respiratory tract showed a PPA of 94.9 % and a NPA of 97.6 % compared to the results of the manual R-GENE RT-PCR. Unfortunately, a precise LOD could not be determined for the PF LDT due to a lack of quantified reference materials but the high amplification efficiencies of the PF LDT indeed suggested low LODs. This may also be supported by our diagnostic experience with the PF LDT in the winter season 2019/2020. 87 of 1732 diagnostic specimens were tested positive for endemic hCoV in the PF LDT. HCoV HKU1 predominated in this season (71 out of 87 coronavirus positive specimens). Other positive specimens were distributed as following: NL63 10 positives, 229E 4 positives and OC43 2 positives. Another limitation of the study was that HKU1 was not included in the panel of proficiency testing specimens probably because HKU1 cannot be grown in cell culture easily ^40^. As the R-GENE assay does not differentiate the endemic hCoV, we could not confirm that a sample positive for HKU1 in the PF LDT truly contains HKU1 RNA but 13 of 15 samples positive for HKU1 were at least confirmed as hCoV positive in the R-GENE assay. Moreover, the primer and probe sequences for HKU1 (and all other hCoVs) that had previously been published ^28^ were reanalyzed on specificity using Nucleotide Blast (NCBI PubMed). As no homology of the primer and probe sequences with another than the intended corresponding hCoV sequence was detected, we presume classification to be correct.

A fast and accurate differentiation of respiratory pathogens in patients with respiratory symptoms is essential to enable sufficient infectious control measures, particularly with regard to the current SARS-CoV-2 pandemic. Together with the PF respiratory panel, 15 different respiratory viruses (Flu A/B, RSV, ParaFlu 1-4, hMPV, AdV, RhV, hCoV NL63/229E/HKU1/OC43 and SARS-CoV-2) can be detected within 4 hours from one single specimen loaded on the PF. Alternatively, a cost saving step by step diagnostic approach is feasible on the PF, e.g. starting with SARS-CoV-2 testing and if negative, testing for other respiratory viruses subsequently.

## Data Availability

Supplmenentary data is included in the manuscript file

## Acknowledgements

We want to thank Prof. Annemarie Berger (Institut für Medizinische Virologie, Universitätsklinikum Frankfurt, Germany) for providing us SARS-CoV-1 cell culture supernatants and Dr. Victor Corman (National Reference Laboratory for Corona Viruses, Charité, Berlin) for providing us hCoV NL63, OC43 and 229E cell culture supernatants.

---

**Supplementary Table 1:**
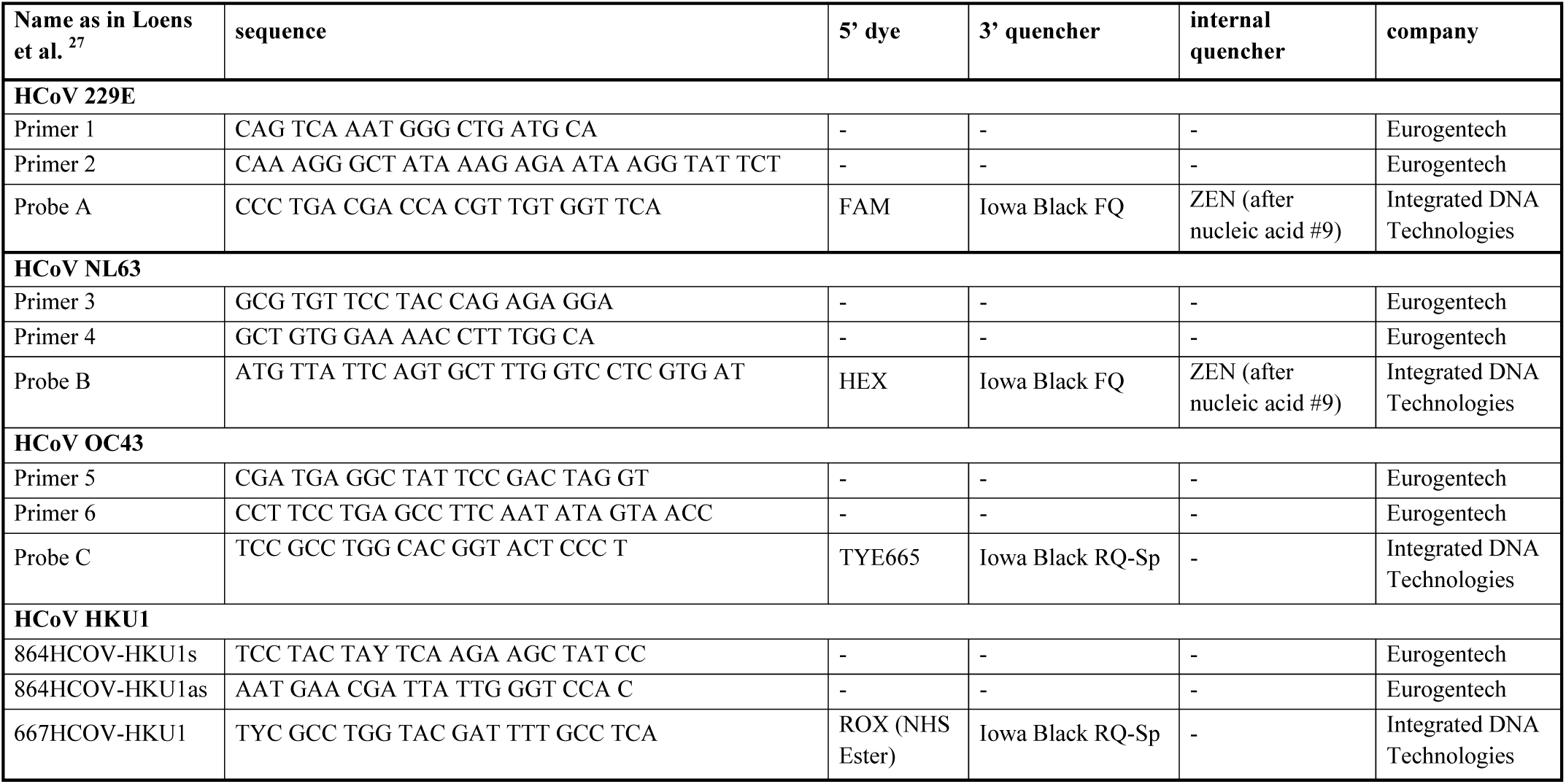
Primer and probe sequences were used as published by Loens et al. Modifications were adapted for multiplexed use as shown in the table. All primers and probes were synthesized HPLC purified.

**Supplementary table 2:** Stability of the PPF on the PF. A PPR was pipetted and loaded onto the PF platform on day 0 and stayed on the platform for fourteen days. The same samples were tested on day 0, 6, 8 and 14.

|  | **Sample** | **material\*** | **Result**<br>**Day 0**<br>**(Ct value)** | **Result**<br>**Day 6**<br>**(Ct value)** | **Result**<br>**Day 8**<br>**(Ct value)** | **Result**<br>**Day 14**<br>**(Ct value)** |
| --- | --- | --- | --- | --- | --- | --- |
| 1 | NC** | BL | negative | negative | negative | negative |
| 2 | NC** | NPS | negative | negative | negative | negative |
| 3 | 229E | NPS | 25.5 | 26.3 | 26.0 | 26.4 |
| 4 | 229E | PL | 34.4 | 34.4 | 33.3 | 33.9 |
| 5 | NL63 | PL | 19.2 | 18.0 | 18.8 | 19.3 |
| 6 | NL63 | BAL | 34.4 | 34.1 | 34.2 | 35.0 |
| 7 | HKU1 | BAL | 26.9 | 25.7 | 27.2 | 28.7 |
| 8 | HKU1 | BAL | 36.6 | 38.1 | 36.5 | 37.5 |
| 9 | OC43 | NPS | 23.9 | 24.3 | 26.3 | 24.5 |
| 10 | OC43 | BAL | 36.9 | 39.4 | 38.7 | 38.0 |

\* bronchial alveolar lavages (BAL), bronchial lavages (BL), nasal swabs (NS), nasopharyngeal swabs (NPS), pharyngeal lavages (PL), tracheal swabs (TRS), tracheal secretions (TS)
\*\* negative control (NC)

